# Predicting Adverse Events in the Cardiothoracic Surgery Intensive Care Unit Using Machine Learning: Results and Challenges

**DOI:** 10.1101/2022.12.16.22283463

**Authors:** Saeed Amal, Robert Kramer, Douglas Sawyer, Jaime B Rabb, Alanna S Maurais, Cathy S. Ross, Alexander Iribarne, Raimond L Winslow

## Abstract

It is highly important to anticipate impending problems in patients in the cardiothoracic intensive care unit (CTICU) and be proactive with respect to prediction of adverse events, enabling interventions to prevent them. In order to develop models that predict the occurrence of adverse events after cardiac surgery, a dataset of 9,237 patients was constructed of a single center’s Society of Thoracic Surgeons (STS) internal database. 1,383 of those patients had developed at least one of seven defined adverse events for this analysis. For the control set, we randomly picked 1,383 patients from the group who did not develop any adverse event. The ensemble learning algorithm, random forest, was applied and outperformed the best reported logistic regression models for similar task (c-statistic of ∼0.81), by achieving an AUC of 0.86 with a 95% CI of [0.81-0.90], specificity of 0.72, sensitivity of 0.82, PPV of 0.78 and NPV of 0.77. In the future, we plan to run a similar evaluation process on a multicenter dataset, and then use this static prediction model as a context for using time-evolving data to develop algorithms for real-time feedback to care teams. In acute care settings, such as the operating room and intensive care unit, the ability to anticipate potentially fatal complications will be enhanced by using supervised machine learning algorithms.

## Introduction

When untoward events occur after cardiac surgery, a rapid response is needed. The intensive care environment offers the patients the best chance of survival. Risk-adjusted mortality rates following cardiac surgery vary 2.5-fold across low and high-performing hospitals [Ghaferi et al. 2009] and a portion of the variability in hospital mortality rates may be attributed to differences in a concept known as “failure to rescue.” Failure to rescue (FTR) is the inability to prevent a patient’s death following a complication. Enhancing the ability to predict or respond rapidly and effectively to untoward events translates into lower morbidity and mortality rates. For example, in Reddy et al. [2013], the following seventeen complications were associated with FTR: Multi-system organ failure, coma, cardiac arrest, renal dialysis, sepsis, anticoagulation event, gastrointestinal event, intensive care unit readmission, prolonged ventilation, reoperation for bleeding, pneumonia, stroke, cardiac tamponade, pulmonary embolism, deep sternal wound infection, heart block, and aortic dissection. At Maine Medical Center (MMC) the FTR metric is tracked on a statistical process control chart that is published quarterly depicting the last 5 years of experience. An example of a statistical process control chart for FTR is shown in Figure 1.

**Figure 1:**
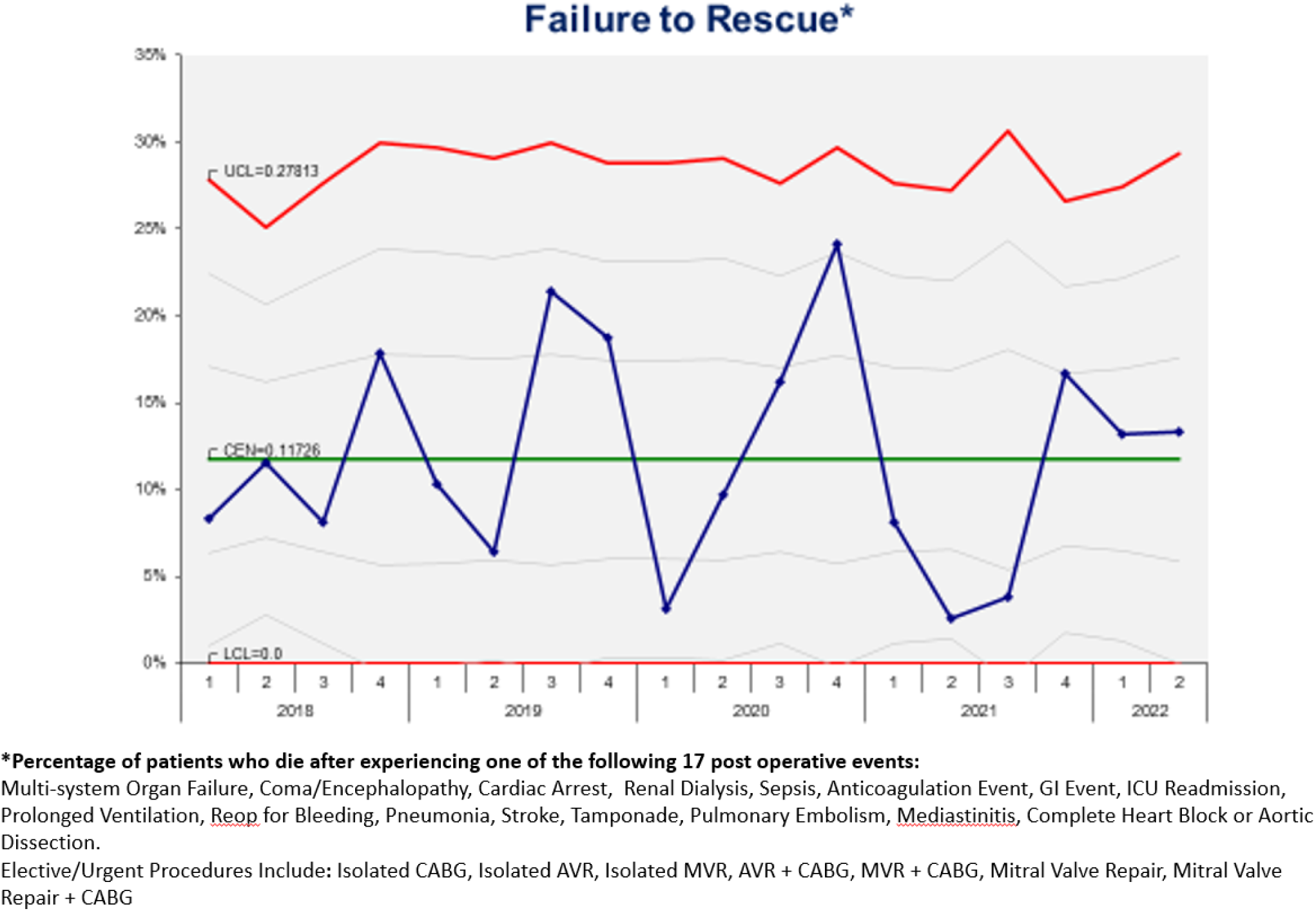
Statistical process control chart for Failure To Rescue.

Using a different set of four variables (prolonged ventilation, stroke, reoperation, and renal failure), Kurlansky et al. [2021], built the binary classification ability of the task of distinguishing between failure to rescue (FTR) cases or not. However, although we did not evaluate on the same data set, their best performing logistic regression analysis [Sperandei 2014] achieved a c-statistic of 0.81 in the binary classification task of identifying FTR cases. Kurlansky did not report additional performance metrics such as sensitivity, specificity, PPV, NPV or 95% CI. Figure 2 demonstrates the AUC graph of logistic regression and random forest respectively. On earlier works using the STS Adult Cardiac Surgery Database, Shahian et al. [2018] and O’Brien et al. [2018] developed risk scores for nine different outcomes of interest: Operative mortality, stroke, renal failure, prolonged ventilation, reoperation, mediastinitis/deep sternal wound infection (DSWI), major morbidity or mortality composite, prolonged postoperative length of stay (POLOS), or short POLOS. O’Brien et al. [2018] identified 70 static variables used to construct risk scores for these outcomes. (Supplement 1)

**Figure 2:**
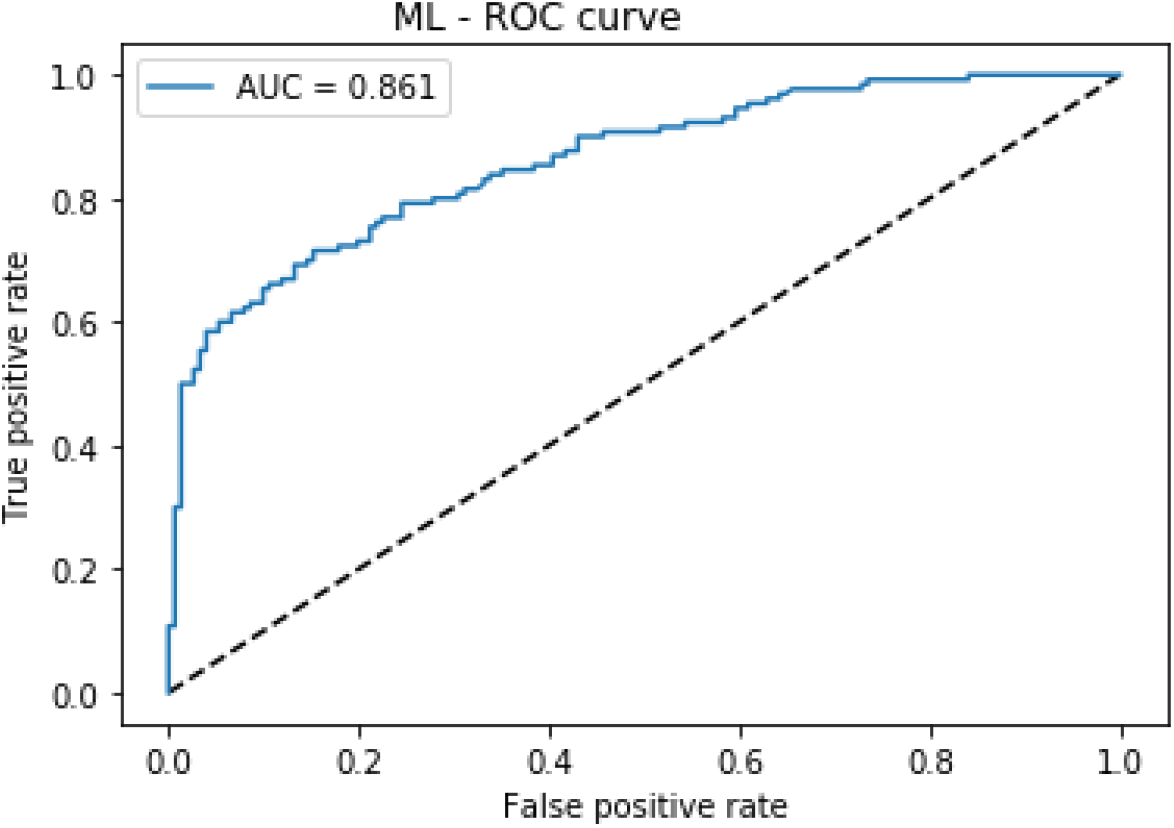
Area under the curve for the random forest model with all adverse events.

The following are shortcomings of the current works quoted above:

1. The results cannot be replicated because the functional form of the risk estimators is not described in either Shahian et al. [2018], O’ Brien et al. [2018], or Kurlansky et al. [2021].
2. The performance of these risk models is not described in a way that supports their full evaluation as predictors.
3. The performance of these risk scores is only assessed by the sole metric provided (the c-statistic, another name for the more commonly used term AUC).
4. The relative weight of each variable used to generate the risk scores (Supplement 1) was not included in the publications.

In our approach, we addressed these limitations by providing insights on the most important variables that affected the predicted outcome. These outcomes are reported through evaluation of the AUC. The curve referred to is the receiver operating characteristic (ROC) curve. In the context of the ROC, sensitivity is the proportion of true cases that are predicted as being true and specificity is the proportion of false cases that are predicted as being false. The positive predictive value (PPV) is the proportion of all true cases that are true, and the negative predictive value (NPV) is the proportion of all false cases that are false.

## Methods

### Data source

The Society of Thoracic Surgeons (STS) Adult Cardiac Surgery Database (ACSD) was queried to develop a dataset including cardiac surgery cases from a single center (Maine Medical Center) over a 9-year period from January 1, 2012 to December 31, 2021, which included 9,237 patients. All patient identifiers and private health information (PHI) were removed for patient protection. The project was submitted to the Maine Medical Center (MMC) Institutional Review Board (IRB), who determined the project to be “non-research” in a letter dated September 11, 2021.

### Cohort

The model for identifying patients who would develop one or more of the following seven adverse events (STS NQF endorsed measures) after cardiac surgery included:

1. Reoperation for any cardiac reason
2. Renal failure
3. Deep sternal wound infection
4. Prolonged ventilation
5. CVA (stroke)
6. 30-day mortality
7. Mortality status at hospital discharge

We searched the 9,237 patient dataset from the MMC STS registry for patients that developed one of the seven adverse events mentioned above, lumped them, and found 1,383 patients, as seen in Table 1. For the control set, we randomly picked 1,383 patients from those who did not develop an adverse event.

**Table 1.** Patient characteristics. *The T-test was used to calculate p-values for group differences in numerical (continuous) variables and chi-squared tests were used to calculate p-values for group differences in categorical variables. Data presented as either mean (SD) or value (%)*.

| **Variable** | **Control cases**<br>**N = 7,854** | **Adverse**<br>**event cases**<br>**N=1,383** | **p-value** |
| --- | --- | --- | --- |
| **Age (SD)** | 64.8 (11.9) | 66.0 (12.3) | 0.0007 |
| **Gender (% Female)** | 2076 (26.4) | 446 (32.2) | <0.0001 |
| **Race (%)** |  |  | 0.7991 |
| *White* | 7704 (98.1) | 1354 (97.9) |  |
| *Black* | 50 (0.6) | 13 (0.9) |  |
| *Asian* | 32 (0.4) | 5 (0.4) |  |
| *Native American* | 19 (0.2) | 2 (0.1) |  |
| *Other* | 49 (0.6) | 9 (0.7) |  |
| **BMI (SD)** | 30.0 (8.9) | 29.3 (6.4) | 0.0009 |
| **Ejection Fraction (SD)** | 55.2 (10.4) | 48.6 (14.7) | <0.0001 |
| **Clinical Status in OR (%)** |  |  | <0.0001 |
| *Non-Urgent* | 3415 (43.5) | 442 (32.0) |  |
| *Urgent* | 4211 (53.6) | 716 (51.8) |  |
| *Emergent* | 227 (2.9) | 218 (15.8) |  |
| *Emergent Salvage* | 1 (0.0) | 7 (0.5) |  |
| **Creatinine (SD)** | 1.04 (0.71) | 1.24 (0.84) | <0.0001 |
| **Hypertension (%)** | 6376 (81.2) | 1122 (81.1) | 0.9625 |
| **Cardiogenic Shock (%)** |  |  | <0.0001 |
| *None* | 7797 (99.3) | 1209 (87.4) |  |
| *At Time of Procedure* | 46 (0.6) | 161 (11.6) |  |
| *<24hr After Procedure* | 11 (0.1) | 13 (0.9) |  |
| **Cardiac Symptoms at Admission (%)** |  |  | <0.0001 |
| *None* | 193 (2.5) | 22 (1.6) |  |
| *Stable Angina* | 241 (3.1) | 14 (1.0) |  |
| *Unstable Angina* | 2358 (30.2) | 195 (14.1) |  |
| *NSTEMI* | 1717 (21.9) | 329 (23.8) |  |
| *STEMI* | 193 (2.5) | 107 (7.7) |  |
| *Angina Equivalent* | 209 (2.7) | 33 (2.4) |  |
| *Other* | 2933 (37.3) | 683 (49.4) |  |

### Existing models

To compare our results to the performance of existing models, we searched and found the models developed by Shahian et al. [2018], O’ Brien et al. [2018], and Kurlansky et al. [2021]. The performance of these risk models in O’Brien et al. are not described in a way that supports their full evaluation as predictors and they are evaluated by only the c-statistic (AUC), which were modest, ranging from 0.57 to 0.81. In O’Brien et al., “…the bootstrap-adjusted c-statistics were lowest for reoperation (range, 0.574 to 0.627) followed by stroke (range, 0.616 to 0.704) and were highest for renal failure (range, 0.749 to 0.810).” AUC of 0.57 is only slightly better than that of a random unbiased coin toss.

### Our model

We built machine learning models using MMC data divided into cases and controls. For the missing values of the data we used the imputation technique of the most common value in the dataset. We used the ensemble learning technique of random forest [Malley et al. 2011]. 90% of the patients in our dataset were used for building a model and 10% were reserved for accuracy testing.

## Results

After creating the cohort of 1,383 patients with at least one of the adverse events and the control group of 1,383 patients that did not have any adverse events, the patients were randomly sampled in order to build a balanced dataset. They were divided into 90% and 10% training and test set respectively. The random forest model achieved an AUC of 0.86 and better specificity of 0.72, which indicates higher ability of identifying patients that will have complications. The model has a sensitivity of 0.82, which indicates a lower ability of identifying patients that will not have complications, a lower performance on PPV (positive predictive value) of 0.78 and NPV (negative predictive value) of 0.77 and higher confidence with a 95% CI of [0.82-0.9]. Figure 2 shows the area under the curve for this model.

In order to understand the input variables that contributed the most to the model outcome, see supplement 1, a list of all variables used as input to the models and supplement 2, a list of the 20 ranked variables that contributed most to outcomes according to the random forest model.

The top five of the 20 variables that most contributed to outcomes are:

1. When IABP was inserted
2. Lowest measured hematocrit recorded in the operating room
3. Ejection Fraction
4. Platelet count closest to the date and time prior to surgery but prior to anesthetic management
5. Patient age

## Data Availability

All data produced in the present study are available upon reasonable request to the authors

## Limitations

While this model is based on a large sample size from a nine-year period, it is a single center experience and a considerably smaller sample size than that upon which the existing logistic regression prediction models that we used for comparison are based. Future models will be based on a larger multi-center dataset.

## Summary

We built a machine learning model using the ensemble learning technique, random forest, which employs multiple learning algorithms. Nine recent years of the STS database were queried at a single center. We created a prediction model that out-performed existing logistic regression prediction models in cardiac surgery as our prediction model had an AUC of 0.86 for multiple adverse events. Furthermore, the dynamic design of this multi-algorithmic machine learning model is designed to improve with time and more data. In the future, combining this static data with time-evolving data in dynamic settings such as the operating room or intensive care units will drive real-time feedback to the care team with potential improvement in failure to rescue rates. In the future, given the importance of the integration of multimodal data for accurate prediction as shown by Amal et al. [2022], we will be combining the current static data with time-evolving data in dynamic settings such as the operating room or intensive care units will drive real-time feedback to the care team with potential improvement in failure to rescue rates. In addition, as done by Ghanzouri et al. [2022] we will add visualization to reflect the risk score with the physicians. Additional future work, we will perform user study similar to Ho et al. [2022] that will collect feedback from the surgeons.

---

**Supplement 1.**
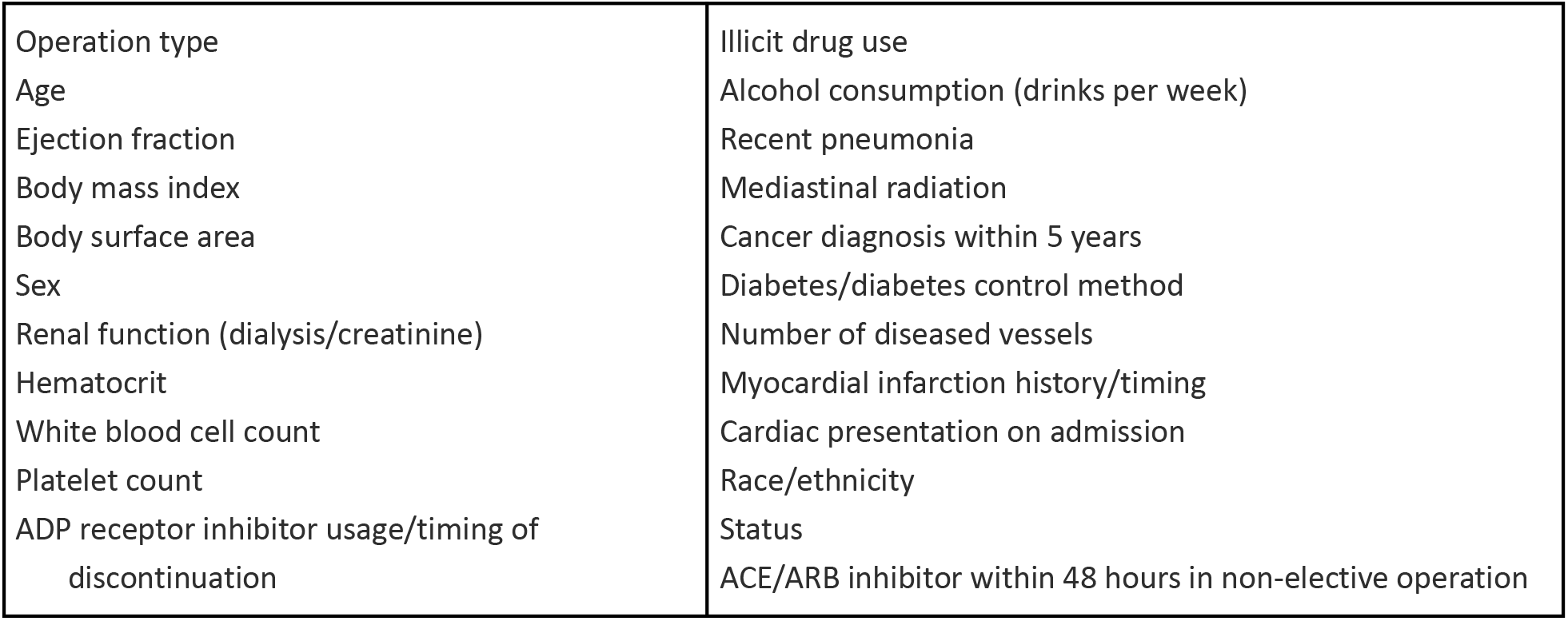
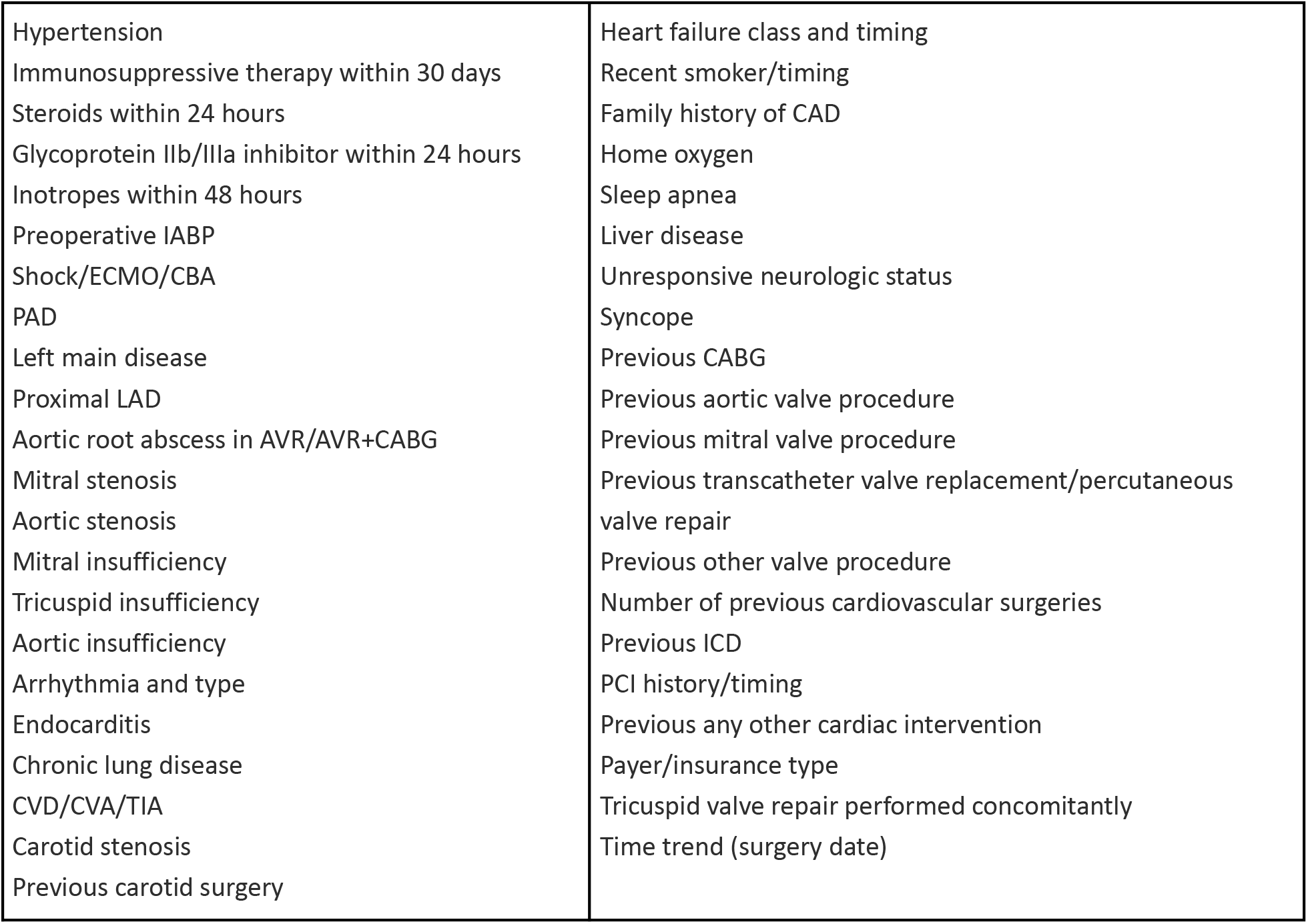
Static variables used to construct risk scores for these outcomes identified by O’Brien et al [2018]. ACE = angiotensin-converting enzyme; ADP = adenosine diphosphate; ARB = angiotensin-receptor blocker; AVR = aortic valve replacement; CABG = coronary artery bypass grafting surgery; CAD = coronary artery disease; CBA = catheterization-based assist device; CVA = cerebrovascular accident; CVD = cardiovascular disease; ECMO = extracorporeal membrane oxygenation; IABP = intra-aortic balloon pump; ICD = implantable cardioverter-defibrillator; LAD = left anterior descending artery; PAD = peripheral arterial disease; PCI = percutaneous coronary intervention; TIA = transient ischemic attack.

**Supplement 2.** Static variables used in random forest algorithm.

|  |  |
| --- | --- |
| Patient Age<br>Pre Op Ejection Fraction<br>Body mass index<br>Height (cm)<br>Weight (kg)<br>Sex<br>Pre Op Dialysis<br>Last Creatinine level<br>Pre Op Hematocrit<br>Pre Op White blood cell count<br>Pre Op Platelet count<br>Pre Op Hypertension<br>Immunosuppressive therapy w/in 30 days of procedure | Pre Op Transient ischemic attack<br>Carotid stenosis<br>Illicit drug use<br>Alcohol Use (drinks per week)<br>Recent pneumonia<br>Pre Op mediastinal radiation<br>Cancer diagnosis within 5 years<br>Pre Op Diabetes<br>diabetes control method<br>Number of diseased vessels<br>Prior myocardial infarction<br>Prior myocardial infarction timing |
| Steroids within 24 hours of procedure | Cardiac presentation on admission |
| Glycoprotein IIb/IIIa inhibitor w/in 24 hours of procedure | Race Documented |
| Inotropes within 48 hours of procedure | Patient Race |
| IABP Insertion Timing | Hispanic, Latino or Spanish ethnicity |
| Pre Op Cardiogenic shock | Operative Status |
| ECMO Insertion Timing | Heart failure history |
| Temp assist Device Type (Open or Catheter) | Heart failure timing |
| Catheter based assist device insertion timing | Heart failure type |
| Peripheral Arterial Disease | NYHA classification |
| Left main coronary artery disease | Pre Op Tobacco use |
| Proximal LAD percent stenosis | Family history of Premature CAD |
| Aortic valve disease etiology | Home oxygen use |
| Mitral valve stenosis | Pre Op Sleep apnea |
| Aortic valve stenosis | Pre Op Liver disease |
| Pre Op Mitral insufficiency grade | Pre Op Unresponsive neurologic status |
| Pre Op Tricuspid insufficiency grade | Pre Op Syncope |
| Pre Op Aortic insufficiency grade | Previous CABG procedure |
| Arrhythmia type | Previous valve procedure |
| Pre Op Endocarditis | Previous valve procedure type |
| Pre Op Chronic lung disease | Previous other cardiac procedure type |
| Pre Op Cerebrovascular disease | Primary payer/insurance type |
| CVA | Tricuspid valve repair performed |
|  | Surgery date |

**Supplement 3.** Top twenty variables with rank of importance.

| Variable name | Rank of Importance |
| --- | --- |
| IABP Insertion Timing | 1 |
| Lowest Intra Op Hematocrit | 2 |
| Pre Op Ejection Fraction | 3 |
| Pre Op Platelets | 4 |
| Patient Age | 5 |
| Height (cm) | 6 |
| Pre Op Tricuspid Regurgitation Grade | 7 |
| Pre Op Mitral Regurgitation Grade | 8 |
| Pre Op NYHA Classification | 9 |
| Operative Status | 10 |
| Number of Diseased Vessels | 11 |
| Primary Coronary Symptom for Surgery | 12 |
| Alcohol use (drinks per week) | 13 |
| Last Creatinine Level | 14 |
| Aortic Valve Stenosis (Y/N) | 15 |
| Proximal LAD Percent Stenosis | 16 |
| Prior myocardial infarction timing | 17 |
| Pre Op Chronic Lung Disease | 18 |
| Home Diabetes Control Method | 19 |
| Pre Op Tobacco Use | 20 |

## Glossary

1. ACE: angiotensin-converting enzyme
2. ADP: adenosine diphosphate
3. ARB: angiotensin-receptor blocker
4. ASCD: Adult Cardiac Surgery Database
5. AUC: Area under the curve
6. AVR: aortic valve replacement
7. CABG: coronary artery bypass grafting surgery
8. CAD: coronary artery disease
9. CBA: Catheterization-based assist device
10. CI: Confidence interval
11. CVA: Cerebrovascular accident
12. CVD: Cardiovascular disease
13. CTICU: Cardiothoracic Intensive Care Unit
14. DSWI: Deep sternal wound infection
15. ECMO: Extracorporeal membrane oxygenation
16. FTR: Failure to rescue
17. IABP: Intra-aortic balloon pump
18. ICD: Implantable cardioverter-defibrillator
19. IRB: Institutional Review Board
20. LAD: Left anterior descending artery
21. MMC: Maine Medical Center
22. NPV: Negative predictive value
23. NQF: National Quality Forum
24. PAD: Peripheral arterial disease
25. PCI: percutaneous coronary intervention
26. POLOS: Postoperative length of stay
27. PPV: Positive predictive value
28. ROC: Receiver operating curve
29. STS: Society of Thoracic Surgeons
30. TIA: Transient ischemic attack

